# Identification of severity zones for mitigation strategy assessment COVID-19 outbreak in Malaysia

**DOI:** 10.1101/2020.05.19.20107359

**Authors:** Tahir Ahmad, Azmirul Ashaari, Siti Rahmah Awang, Siti Salwana Mamat, Wan Munirah Wan Mohamad, Amirul Aizad Ahmad Fuad, Nurfarhana Hassan

**Affiliations:** Department of Mathematical Sciences, Faculty of Science, Universiti Teknologi Malaysia, 81310 Skudai, Johor, Malaysia.; Azman Hashim International Business School, Universiti Teknologi Malaysia, 81310 Skudai, Johor, Malaysia.; Department of Mathematics, Faculty of Computer and Mathematical Sciences, Universiti Teknologi MARA, Johor Branch, Pasir Gudang Campus, Jalan Purnama, Bandar Seri Alam, 81750 Masai, Johor, Malaysia

## Abstract

The objective of this research is to identify severity zones for the COVID-19 outbreak in Malaysia. The technique employed for the purpose is fuzzy graph that can accommodate scarcity, quantity, and availability of data set. Two published sets of data by the Ministry of Health of Malaysia are used to implement the technique. The obtained results can offer descriptive insight, reflection, assessment, and strategizing actions in combating the pandemic.

## Introduction

The public panic and discomfort on the ongoing COVID-19 outbreak remind us of the history of the 1918 Spanish Flu pandemic, whereby over 50 million people died worldwide. It was a deadly pandemic, indeed. The ongoing outbreak of coronavirus disease 2019 (COVID-19) has claimed 259 593 lives worldwide as of 8^th^ May 2020, 08:00 GMT, according to the World Health Organization (WHO). Since the first case of pneumonia of unknown cause detected in Wuhan reported to the WHO Country Office in China on 31^st^ December 2019 and followed by its declaration as a Public Health Emergency by the international body on 30^th^ January 2020, researchers, scientists, and mathematicians have been racing in their efforts to stop the potential devastating assault by the coronavirus.

Zhou et al. [1] first tipped off the world to the menace of the virus through their publication in Nature. However, the researchers did not employ any specific mathematical tools in their work. Most mathematical modelers have employed Ordinary Differential Equation (ODE), such as Liang [2] as a tool in their predictive modeling of COVID-19. Similarly, Qianying et al. [3] adopted the system of ordinary differential equations that previously used to model the pandemic 1918 Spanish Flu for describing the current COVID-19 outbreak. Recently, Krantz and Rao [4] described underreporting cases of COVID-19 for several countries using coupled ODE-wavelets model. There are more than 25 papers and preprints in the literature on ODE or ODE coupled with other methods as 2^nd^ May 2020 to model COVID-19 and related issues.

For instance, Hamzah et al. [5] and Prem et al. [6] utilized a system of ordinary differential equations in their Susceptible-Exposed-Infected-Removed (SEIR) models and Jia et al. [7] modified SEIR model for the purposes.

However, to the best of our knowledge, there is only a publication that used graph for COVID-19 related issues currently. It is in Forster et al. [8] whereby the researchers analyzed the coronavirus genomes using the phylogenetic network, a special type of graph that has been used mainly in archeological studies.

There are three main downsides for modeling COVID-19, namely, scarcity, quantity, and availability of data that are essential to produce a good reliable mathematical model. This is due to the fact that the outbreak is about six months old since the first case was reported. Therefore, a flexible and robust mathematical technique that can handle such identified shortcomings is necessary to model the outbreak. In this paper, a fuzzy graph analysis method is presented, namely fuzzy autocatalytic set, that is capable of accommodating such constraints to analyze the current pandemic.

## Methods

Generally, a graph represents a relationship between objects. Objects are represented as vertices and the relations by edges. A graph is formally defined as the following.

**Definition 1** (see [9]). A graph is a pair of sets (*V,E*) where *V* is the set of vertices and *E* is the set of edges.

Furthermore, another way to represent a graph is by its adjacency matrix. The definition of an adjacency matrix for a graph is given in Definition 2 below.

**Definition 2** (see [9]). An adjacency matrix of graph *G*(*V, E*) with *n* vertices is an *n×n* matrix denoted by *A*(*a_ij_*), where *a_ij_* = 1 if *E* contains a directed edge (*j, i*). It is an arrow pointing from vertex *j* to vertex *i*, and *a_ij_ =* 0 otherwise.

**Figure 0.1.**
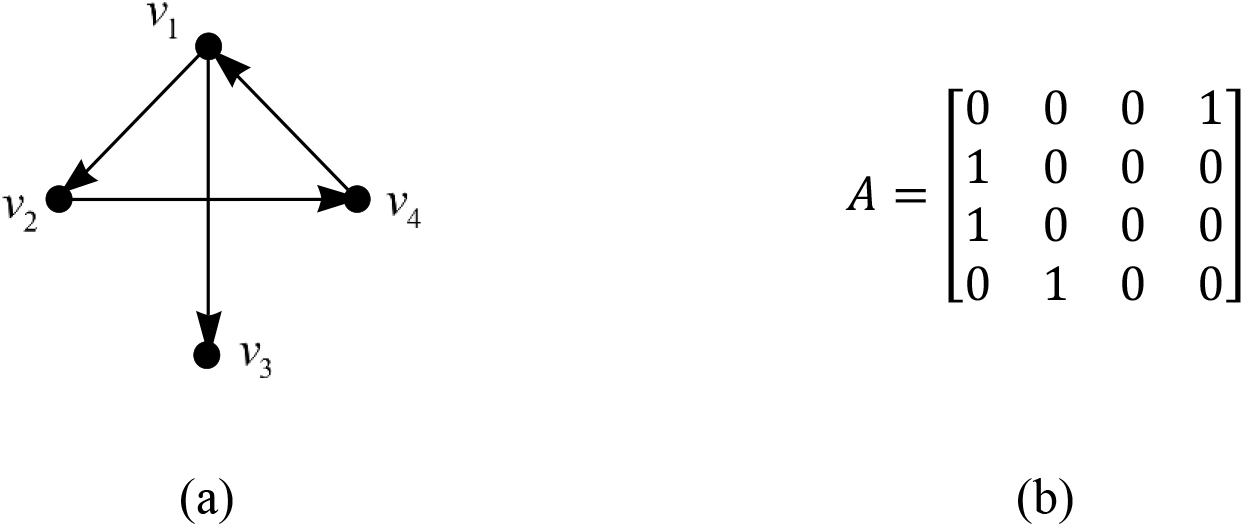
A directed graph (a) and its adjacency matrix (b).

### Fuzzy Autocatalytic Set

The concepts of graph and fuzzy set have given ‘birth’ to a new mathematical structure, namely, fuzzy graph. Definition 3 indicates that vertices and edges are both fuzzy. In other words, the vertices and edges have values between 0 and 1. Figure 0.2 illustrates a fuzzy graph.

**Definition 3** (see [10]). A fuzzy graph *G*(*σ,µ*) is a pair of function *σ:S* → [0,1] and *µ:S × S* → [0,1] such that ∀*x*, *y* ∈ *S, µ*(*x, y*) *≤ σ*(*x*) Λ σ(*y*).

An adjacency matrix of a fuzzy graph is defined as follows:

**Definition 4** (see [10]). An adjacency matrix, *A* of a fuzzy graph *G =* (*V,σ,µ*) is an *n×n* matrix defined as *A =* (*a_ij_*) such that *a_ij_ = µ*(*v_j_, v_i_*).

**Figure 2.**
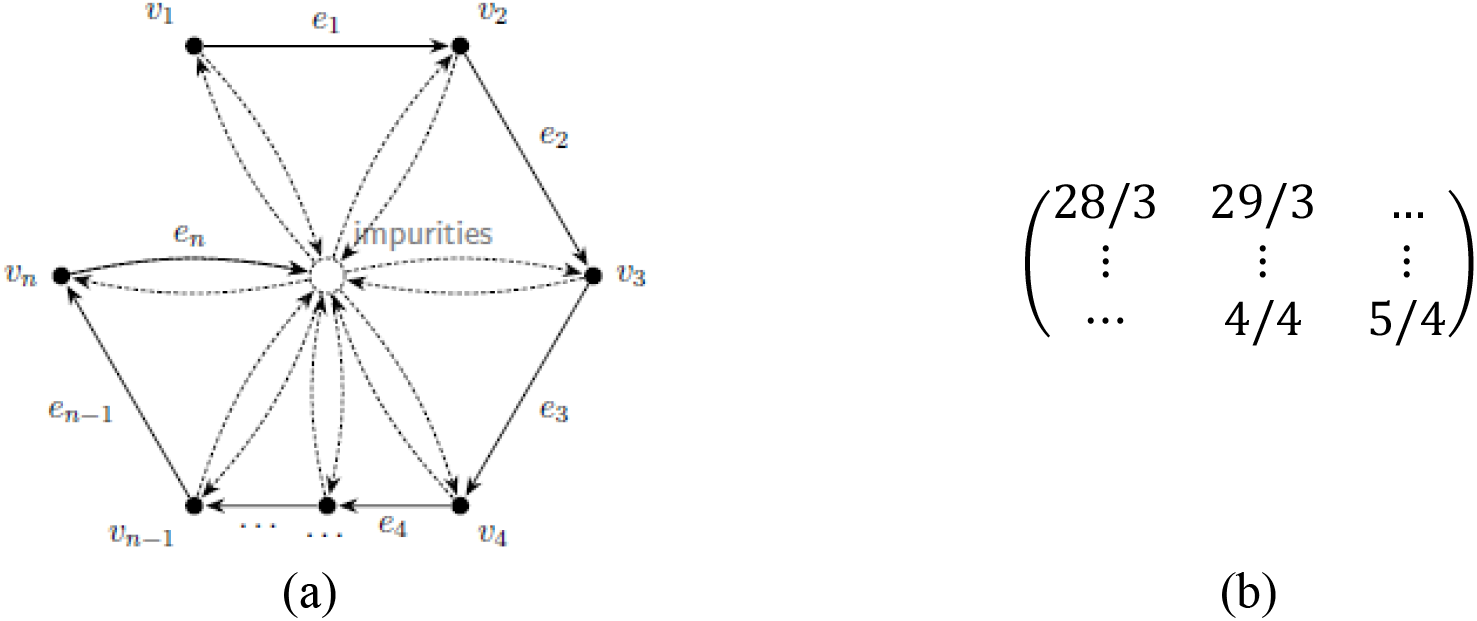
(a) A fuzzy graph and its (b) adjacency matrix with new cases as entries with respect to the data from 28^th^ March to 5^th^ April 2020.

The concept of autocatalysis was originated in chemistry, in particular, for the description of catalytic interaction between molecules [11], [12]. Further, Jain and Krishna [13] formalized the concept of an autocatalytic set (ACS) as a directed graph in which its vertices represent species and edges represent catalytic interactions among them. The formal definition of an ACS is given as follows.

**Definition 5** (see [13]). An autocatalytic set is a subgraph, each of whose vertices has at least one incoming link from vertices belonging to the same subgraph.

Some examples of ACSs are illustrated in Figure 0.3. The simplest ACS is a vertex with 1-cycle.

**Figure 0.3.**
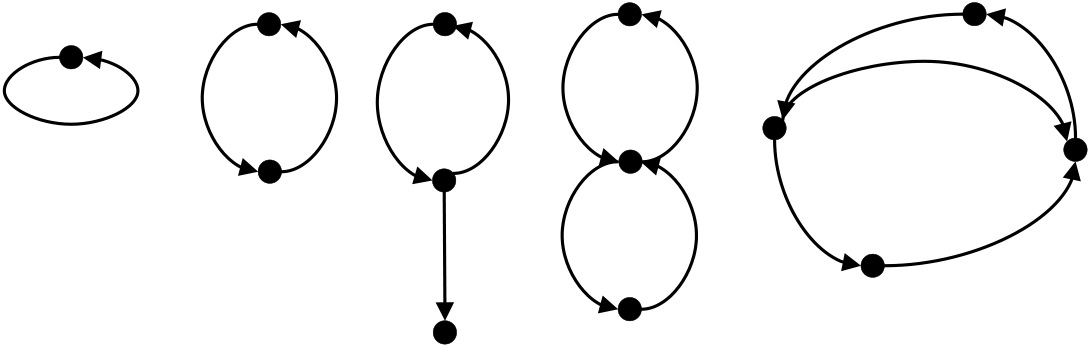
Some examples of ACS

The merger of the fuzzy graph and autocatalytic set has led to the idea of the fuzzy autocatalytic set (FACS) by Tahir et al. [14]. The concept of FACS COVID-19 outbreak in Malaysia is depicted in Figure 0.4. The formal definition of FACS is laid as follows.

**Figure 0.4.**
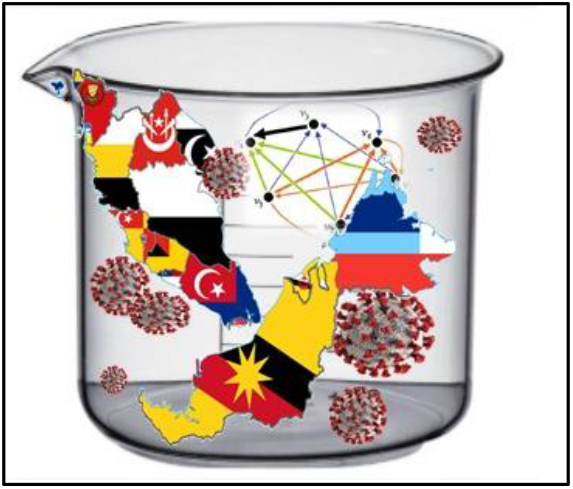
Fuzzy Autocatalytic Set of COVID-19 outbreak in Malaysia.

**Definition 6** (see [14]). A fuzzy autocatalytic set is a subgraph each of whose vertices has at least one incoming link with membership value, *µ*(*e_i_*) ∈ (0,1], ∀*e_i_ ∈ E* from any other vertices are belonging to the same subgraph.

### Dynamics of FACS

The adjacency matrix in Figure 0.1(b) and Figure 0.2(b) is then processed by the procedure outlined in [14], [15] and improved by [16], respectively. The outcomes of the process are determined via the following steps.

Step 1: Keeping *C*(*s × s*) matrix fixed, *x* evolved according to the following equation.

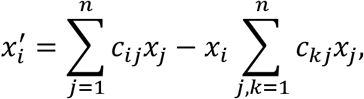

for time *t*, which is large enough for *x* to get reasonably close to its attractor ***X*** (Perron Frobenius Eigenvector). We denoted *X_i_ = x_i_*(*t*).

Step 2: The set *L* of nodes *i* with the least value of *X_i_* is determined, i.e.

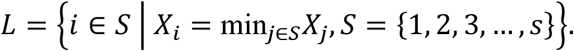

This is the set of “least fit” nodes, identifying the relative concentration of a variable in the attractor (or, more specifically, at *t*) with its “fitness” in the environment defined by the graph. The least fit node is removed from the system along with its links, leading a graph of *s* – 1 variables.

Step 3: *C* is now reduced to (*s –* 1) × (*s –* 1) matrix. The remaining nodes and links of *C* remained unchanged. All these *x_i_*(0 ≤ *x_i_* ≤ 1) are rescaled to keep

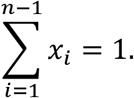

Repeat all the steps until the 2 × 2 matrix is attained.

Figure 0.5 illustrates the initial step (Step 1). Then one of the nodes with the least eigenvector is removed from the graph (Step 2). The node is removed along with its links, and the graph is left with a reduced number of nodes and links (Step 3). This process is then repeated until a graph with at least two nodes is attained.

**Figure 0.5.**
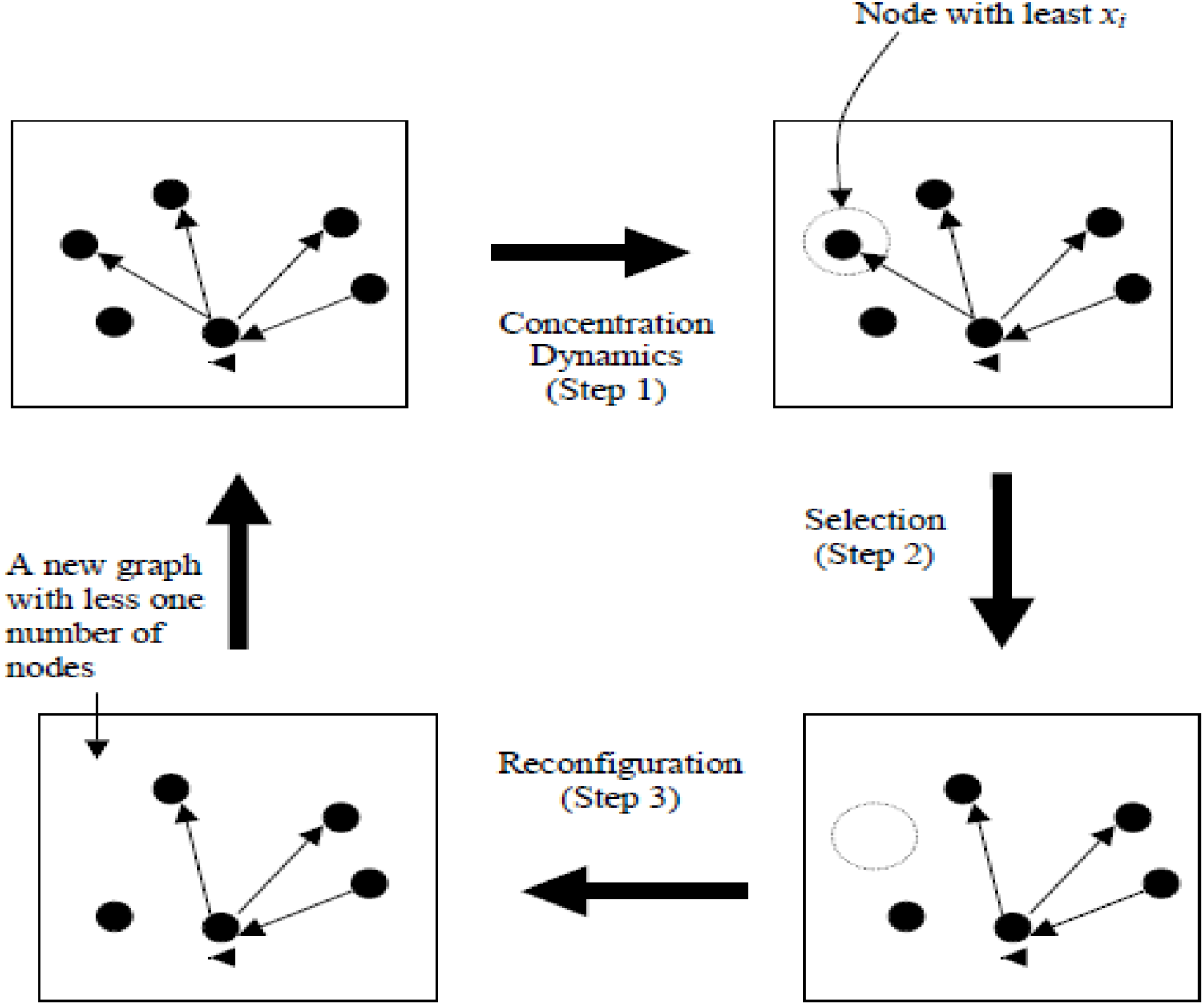
Schematic portrayal of the graph dynamics.

The procedure to transform the graph into 2D-Euclidean space is adopted from [17], which is based on the Laplacian matrix and solving a unique one-dimensional optimization problem in order to determine their coordinates. The general overview of the transformation is depicted in Figure 0.6.

**Figure 0.6.**
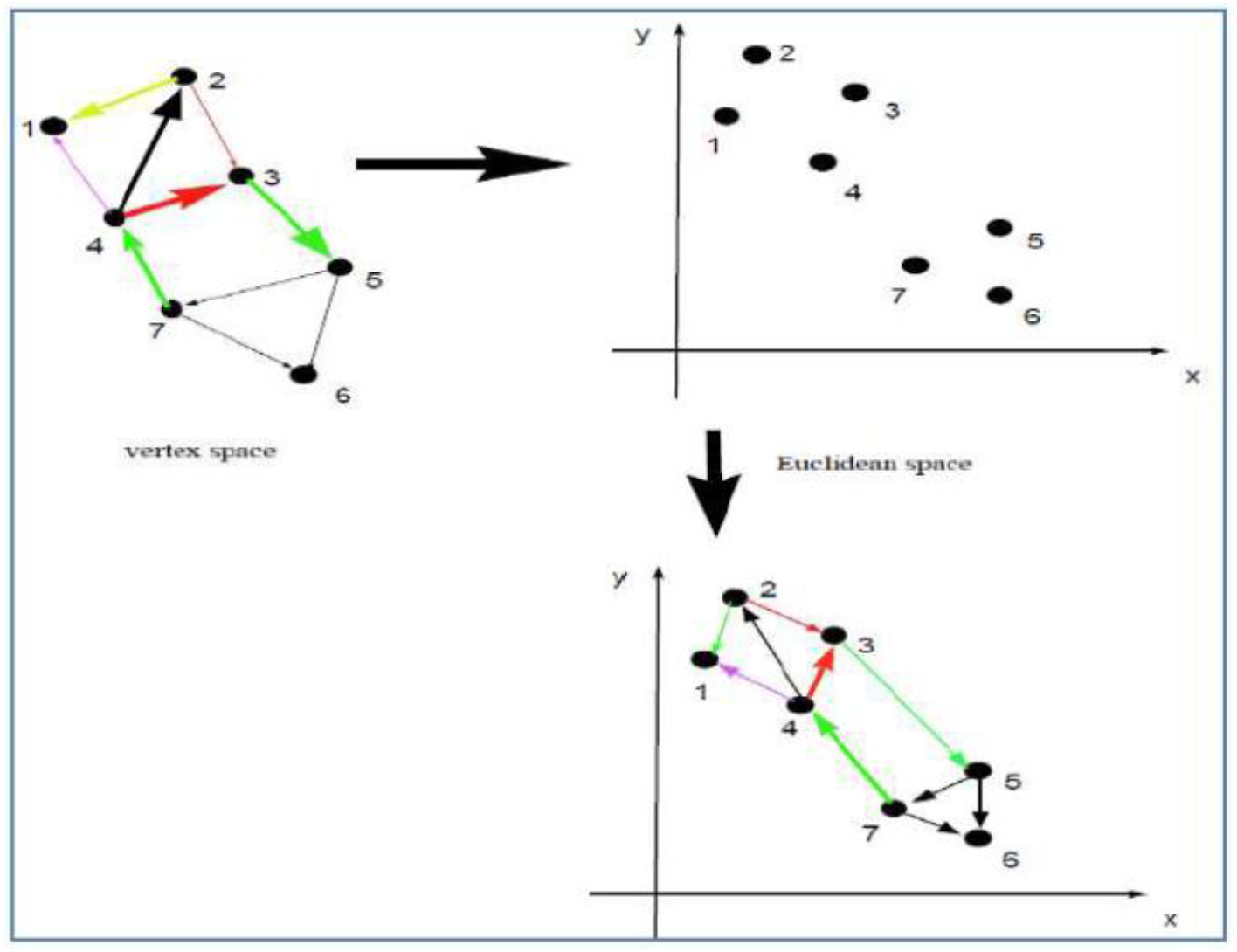
Schematic illustration transformation of the graph from vertex space to Euclidean space.

### Implementation

The proposed technique is implemented on two sets of data. The data are collected from the Ministry of Health, Malaysia (moh.gov.my), and presented in the following subsections. The data of new, death, and recovery cases in Malaysia from 10^th^ March to 10^th^ April 2020 is given in Table 0.1. However, the breakdown of reported new cases between states in Malaysia from 28^th^ March to 5^th^ April is considered for the first case. The period is selected due to the erraticness of the data, as depicted in Figure 0.7a.

**Table 0.1.** New cases in states of Malaysia from 28^th^ March 2020 – 5^th^ April 2020.

| V | State | 28-Mar | 29-Mar | 30-Mar | 31-Mar | 1-Apr | 2-Apr | 3-Apr | 4-Apr | 5-Apr |
| --- | --- | --- | --- | --- | --- | --- | --- | --- | --- | --- |
| 1 | Perlis | 0 | 0 | 1 | 1 | 0 | 0 | 0 | 0 | 1 |
| 2 | Kedah | 1 | 1 | 2 | 0 | 2 | 1 | 1 | 0 | 1 |
| 3 | Pulau Pinang | 6 | 1 | 1 | 6 | 2 | 2 | 0 | 1 | 2 |
| 4 | Perak | 6 | 11 | 8 | 5 | 5 | 6 | 5 | 2 | 6 |
| 5 | Selangor | 33 | 33 | 32 | 32 | 22 | 74 | 63 | 27 | 53 |
| 6 | Negeri Sembilan | 15 | 9 | 10 | 9 | 8 | 18 | 4 | 9 | 11 |
| 7 | Melaka | 9 | 1 | 7 | 2 | 11 | 7 | 3 | 2 | 14 |
| 8 | Johor | 26 | 24 | 24 | 16 | 19 | 27 | 21 | 6 | 16 |
| 9 | Pahang | 14 | 12 | 3 | 3 | 9 | 3 | 13 | 24 | 7 |
| 10 | Terengganu | 4 | 2 | 0 | 0 | 1 | 2 | 18 | 6 | 10 |
| 11 | Kelantan | 10 | 15 | 4 | 4 | 3 | 3 | 0 | 2 | 2 |
| 12 | Sabah | 15 | 0 | 4 | 5 | 3 | 1 | 5 | 10 | 13 |
| 13 | Sarawak | 8 | 11 | 6 | 21 | 32 | 23 | 28 | 7 | 15 |
| 14 | WP Kuala Lumpur | 7 | 28 | 24 | 34 | 25 | 33 | 55 | 52 | 27 |
| 15 | WP Labuan | 5 | 0 | 0 | 0 | 0 | 0 | 0 | 1 | 0 |
| 16 | WP Putrajaya | 0 | 2 | 2 | 2 | 0 | 8 | 1 | 1 | 1 |

**Figure 0.7.**
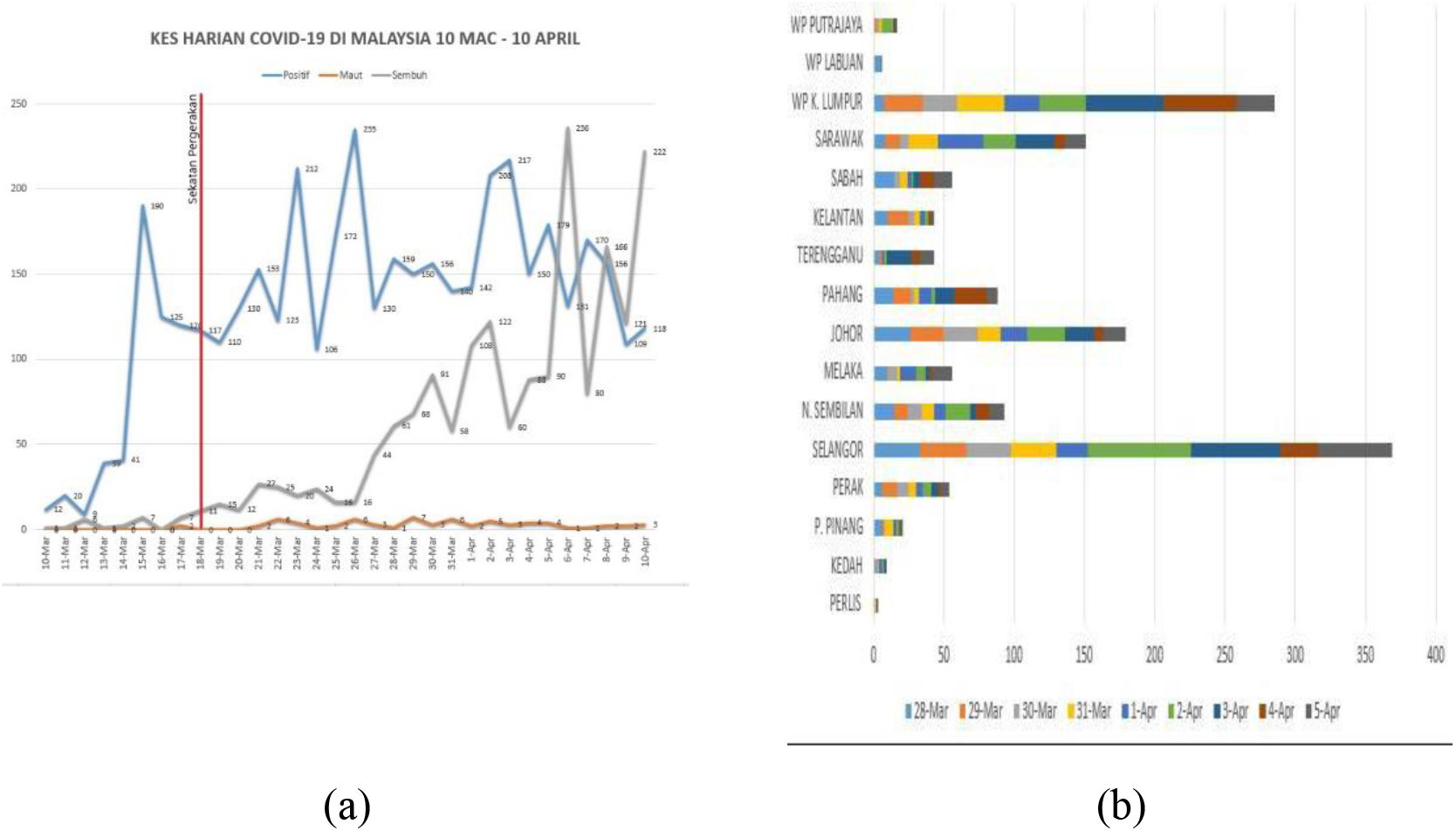
(a) Reported new, death and recovery cases in Malaysia from 10^th^ March to 10^th^ April 2020 (b) New cases in states of Malaysia from 28^th^ March to 5^th^ April 2020.

The data 28^th^ March 2020 – 5^th^ April 2020

The data of new cases of COVID-19 is tabulated in Table 3.1.

The reported new cases in states of Malaysia from 28^th^ March to 5^th^ April 2020 are depicted in Figure 3.1 b, and its adjacency matrix is given in Figure 0.2b.(b)

Using FACS for sampled data 28^th^ March to 5^th^ April 2020, 16 states are identified and clustered, namely, **Cluster 1** contains Perlis, Kedah, Pulau Pinang and Perak. **Cluster 2** includes Selangor, Negeri Sembilan, Melaka and Johor whereas **Cluster 3** is made of Pahang, Trengganu, Kelantan and Sabah. Finally, Sarawak, WP Kuala Lumpur, WP Labuan and WP Putrajaya formed **Cluster 4**.

These clusters are then classified into three zones (refer to Figure 0.8); Zones 1, 2, and 3. Zone 1 is named as the **Under Control** zone that comprises of Perlis, Kedah, Pulau Pinang, and Perak. These four states are scattered in Zone 1, which reflects their disparity with low reported new cases.

**Figure 0.8.**
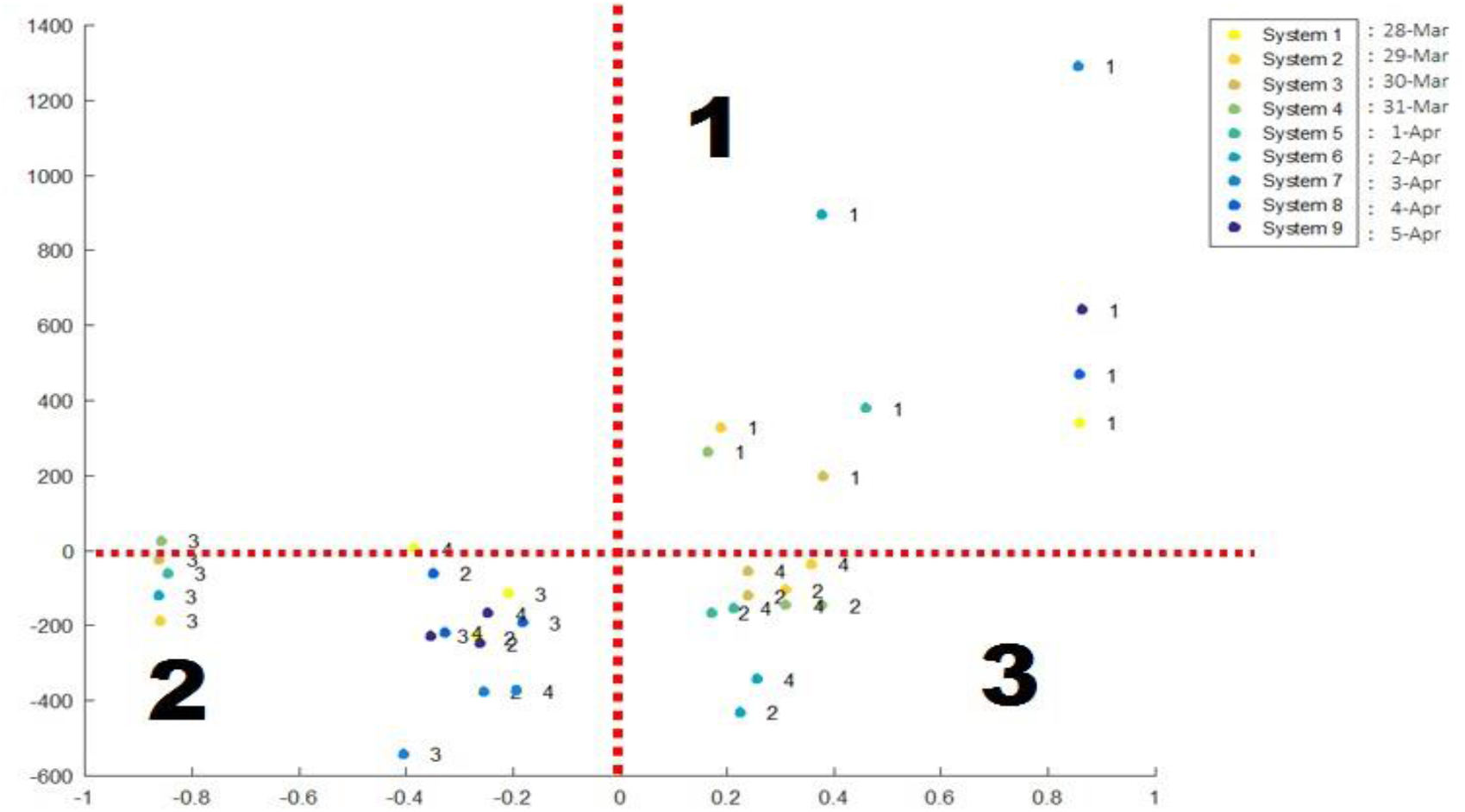
Two phases of FACS clustering for states in Malaysia from 28^th^ Mac to 5^th^ April 2020.

Zone 2 in the **Medium** zone is consisting of Pahang, Terengganu, Kelantan, and Sabah. Increased new cases reported in these states only happened after 31^st^ March. Although Zone 2 is dominated by Cluster 3, it is not total domination since there were a couple of instances where Cluster 2 and 4 popped up in the zone. Hence, the government has to pay attention to the states in Cluster 3 because these states have the potential to move into Zone 3. On top of that, Zone 2 is clearly closed adjacent to Zone 3.

Zon 3 is the **Danger Zone** that is totally dominated by states in the west and south of Malaysia such as Selangor, Negeri Sembilan, Melaka, Johor, WP Kuala Lumpur, WP Putrajaya, including Sarawak and WP Labuan (refer to Figure 0.9).

**Figure 0.9.**
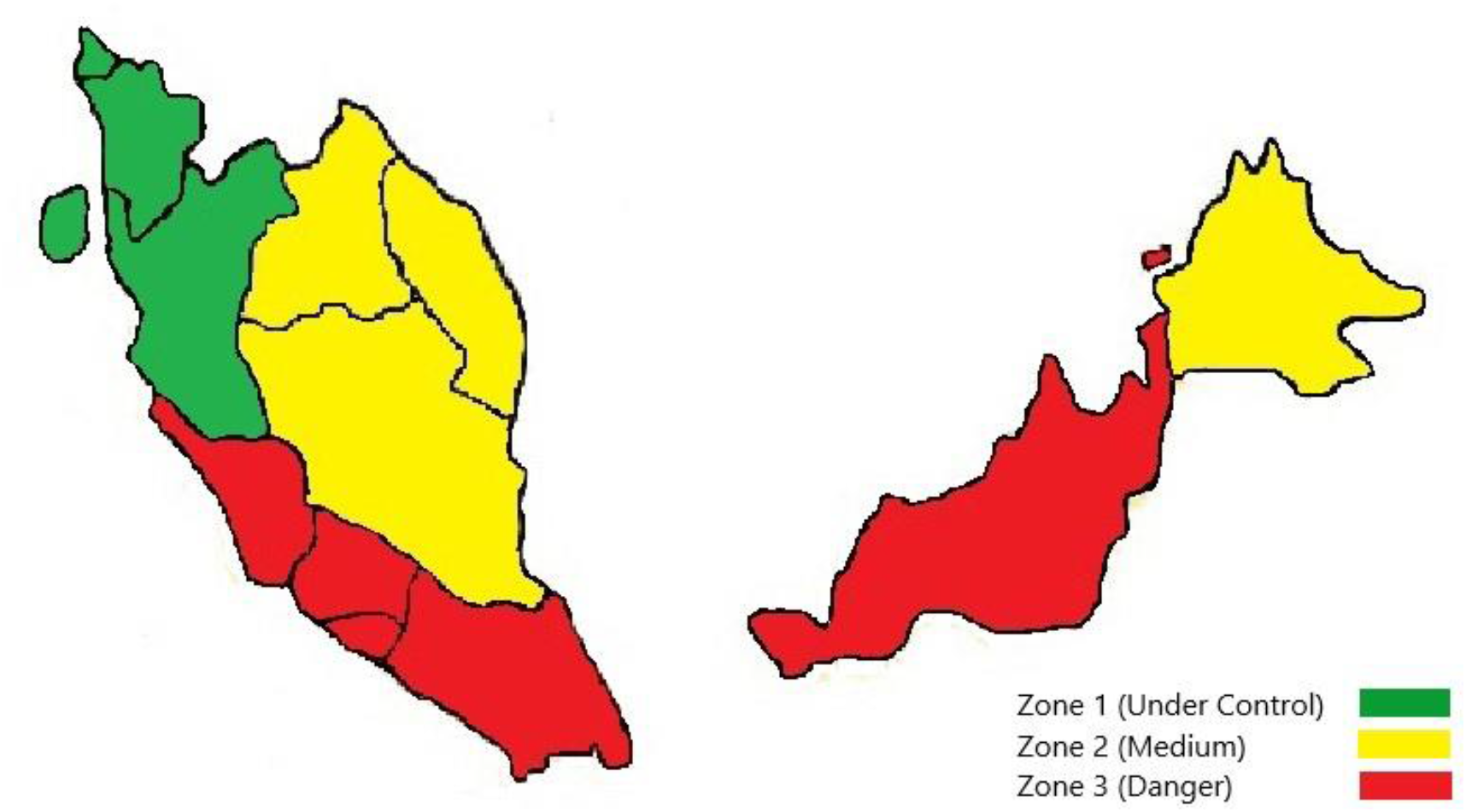
Three Zones with respect to the severity of COVID-19 in Malaysia using FACS.

In fact, the government has gazetted 23 districts in these states as the red zone, namely, Putrajaya, Jasin, Negeri Sembilan, Hulu Langat, Petaling, Johor Bahru, Kuching, and Tawau. The district of Hulu Selangor in the state of Selangor has announced another red zone on 10^th^ April (Figure 0.10a). Our FACS analysis concurred with the list of states in red zones released by Crisis Preparedness and Response Centre (CPRC), Ministry of Health of Malaysia.

**Figure 10.**
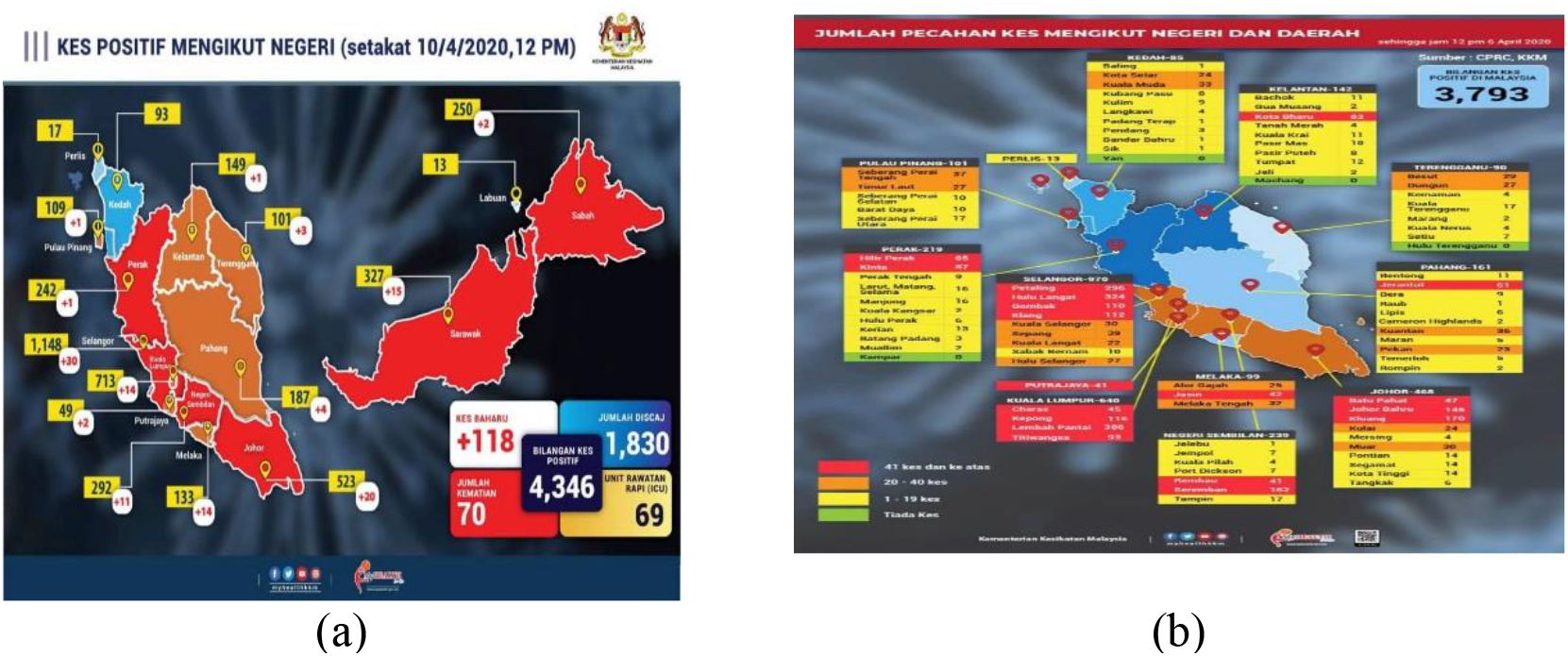
(a) MOH releasedon 10^th^ April 2020 (b) MOH release on 6^th^ April 2020.

Furthermore, we have predicted that WP Putrajaya is in Zone 3 with respect to the data up to 5^th^ April. True enough, WP Putrajaya was announced in the danger zone on 6^th^ April by the government (refer to Figure 0.10b).

### The data 6^th^ April 2020 – 2^nd^ May 2020

The data of new cases of COVID-19 for 6^th^ April 2020 to 2^nd^ May 2020 is tabulated in the attached Appendix. The ordinary graph and bar chart for the data are depicted in Figure 0.11a and Figure 0.11b, respectively.

**Figure 0.11.**
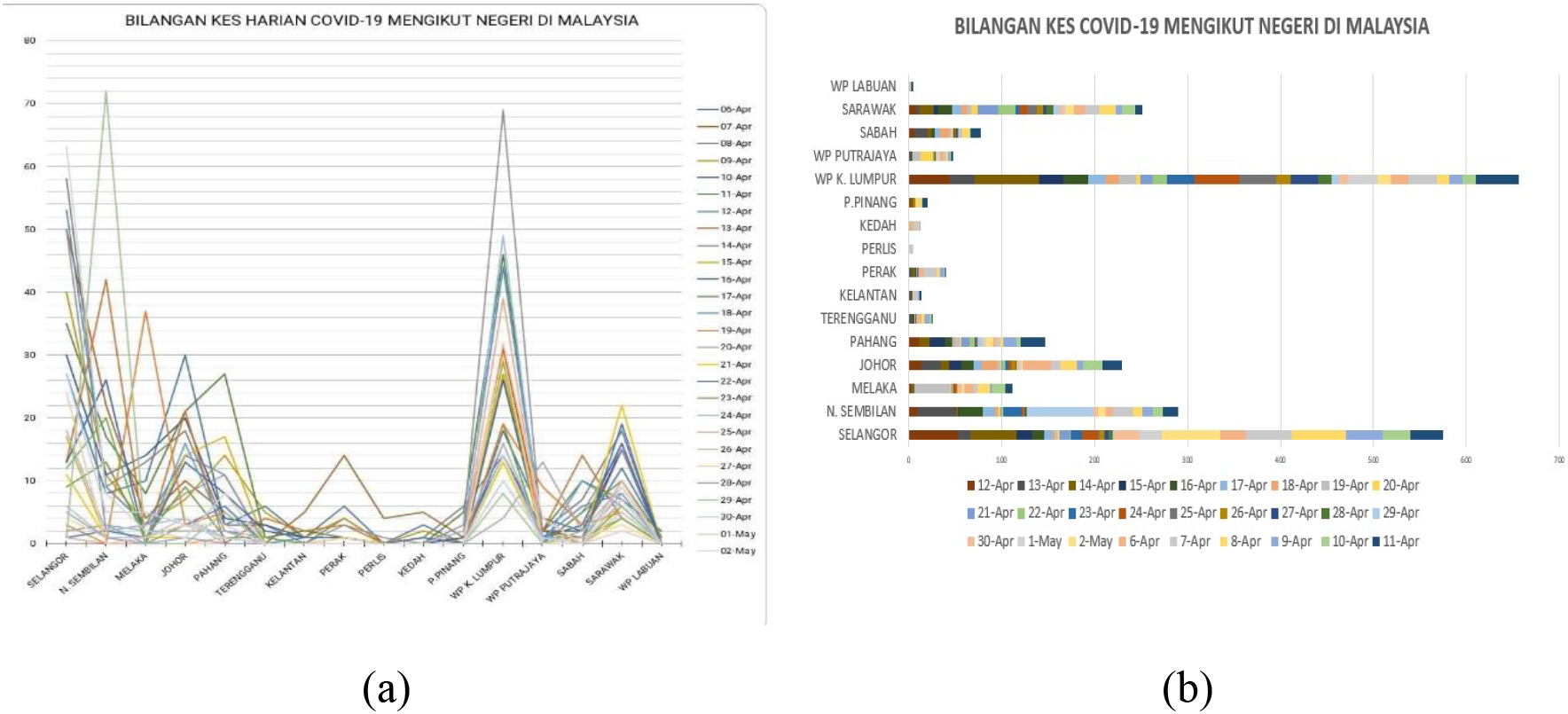
(a) Graph (b) Bar chart for new cases of COVID-19 in Malaysia from for 6^th^ April 2020 to 2^nd^ May 2020

The FACS is then executed and identified four clusters or nodes of states in Malaysia.

**Table 0.2.** Cluster of states in Malaysia.

| Nodes | States |
| --- | --- |
| 1 | Perlis, Kedah, Pulau Pinang, Perak |
| 2 | Selangor, Negeri Sembilan, Melaka, Johor |
| 3 | Pahang, Terengganu, Kelantan, Sabah |
| 4 | Sarawak, WP Kuala Lumpur, WP Labuan, WP Putrajaya |

The coordinates of these nodes on the ***xy*** –plane are determined using coordinated FACS technique and presented in Figure 0.12.

**Figure 0.12.**
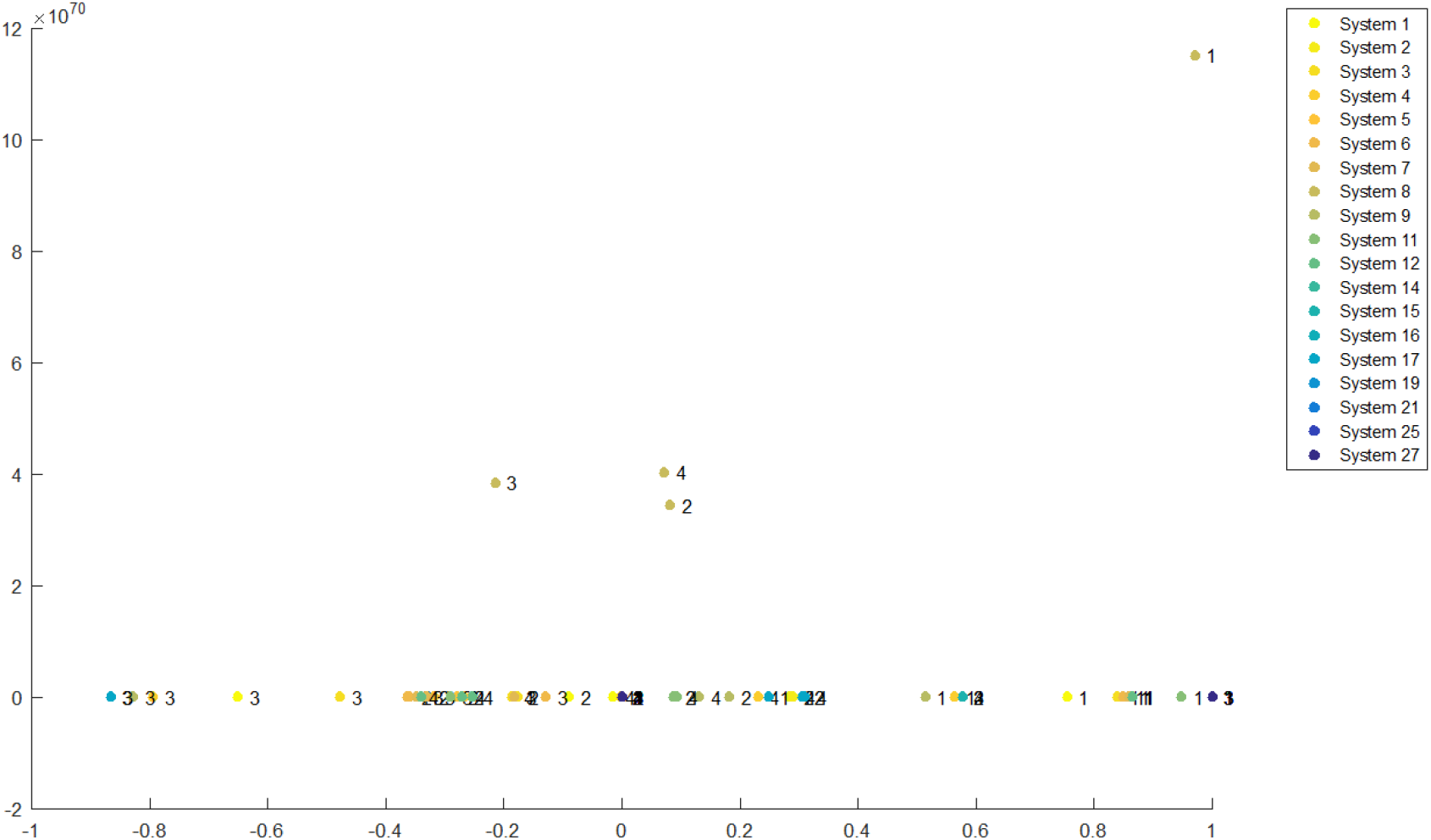
FACS coordinated for new cases 6^th^ April 2020 to 2^nd^ May 2020.

In Figure 0.12, we observed two significant features. First, the majority of the nodes are lined-up at ***y*** *=* **0** (see Figure 0.13a). This feature hints that all the states in Malaysia are responding well with Malaysia’s mitigation strategy during the period. In other words, no states are left behind. Second, there are two axes, namely ***x = –* 0.4** and ***x* = 0.4**, that divide the plane (see Figure 0.13b).

**Figure 0.13.**
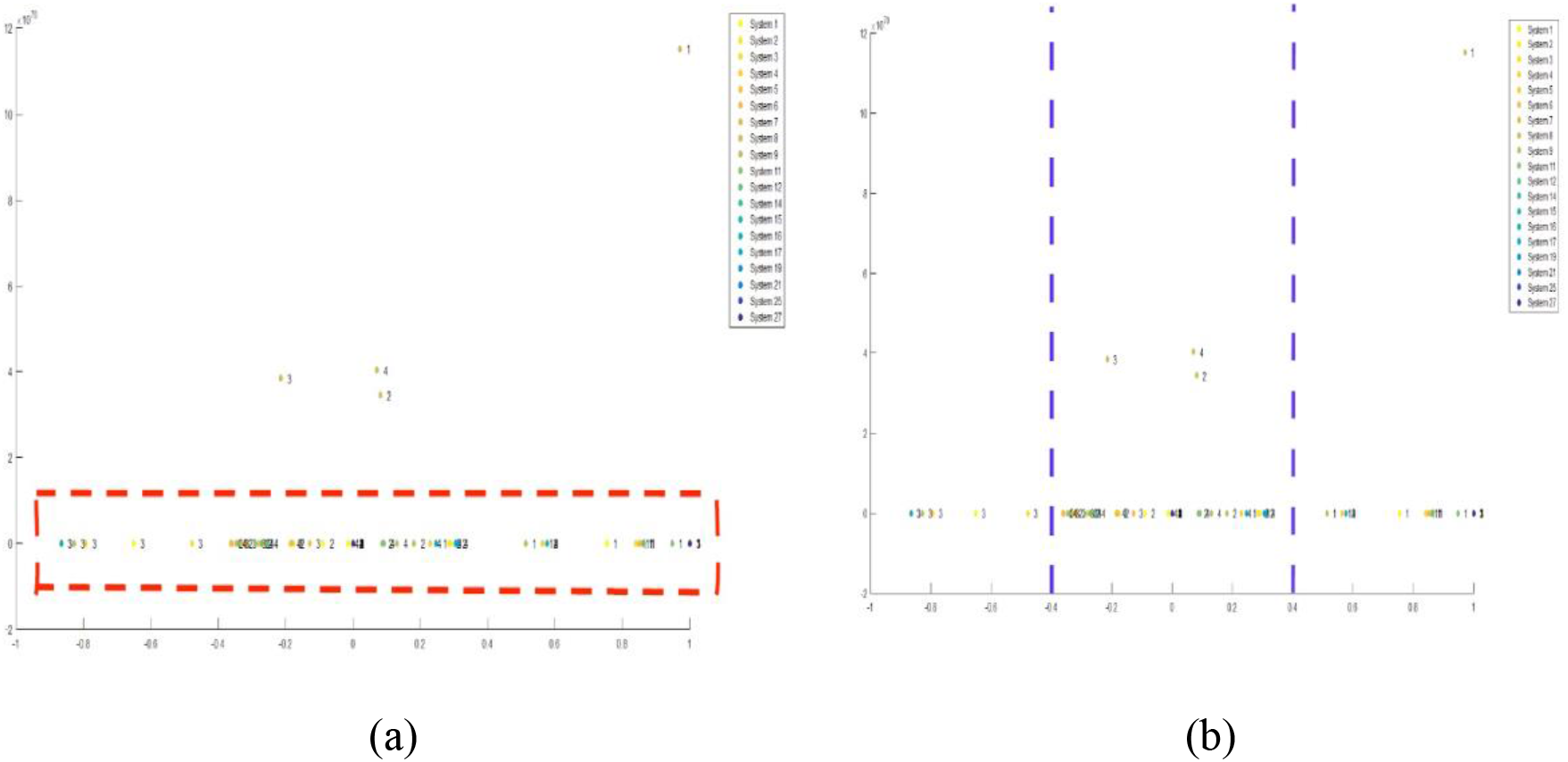
(a) Majority of nodes line up at y = 0 (b) Plane dividers at x = - 0.4 and x = 0.4 Hence, three zones are observed, namely Zone 1 (green), Zone 2 (yellow), and Zone 3 (blue). Zone 1 is dominated by Node 1, Zone 2 by Nodes 2 and 4, and Zone 3 by Node 3. Zone 1 is the most comfortable zone, Zone 2 is where most activities occur, and states in Zone 3 are maintaining their efforts but not out of the wood yet.

**Figure 0.14.**
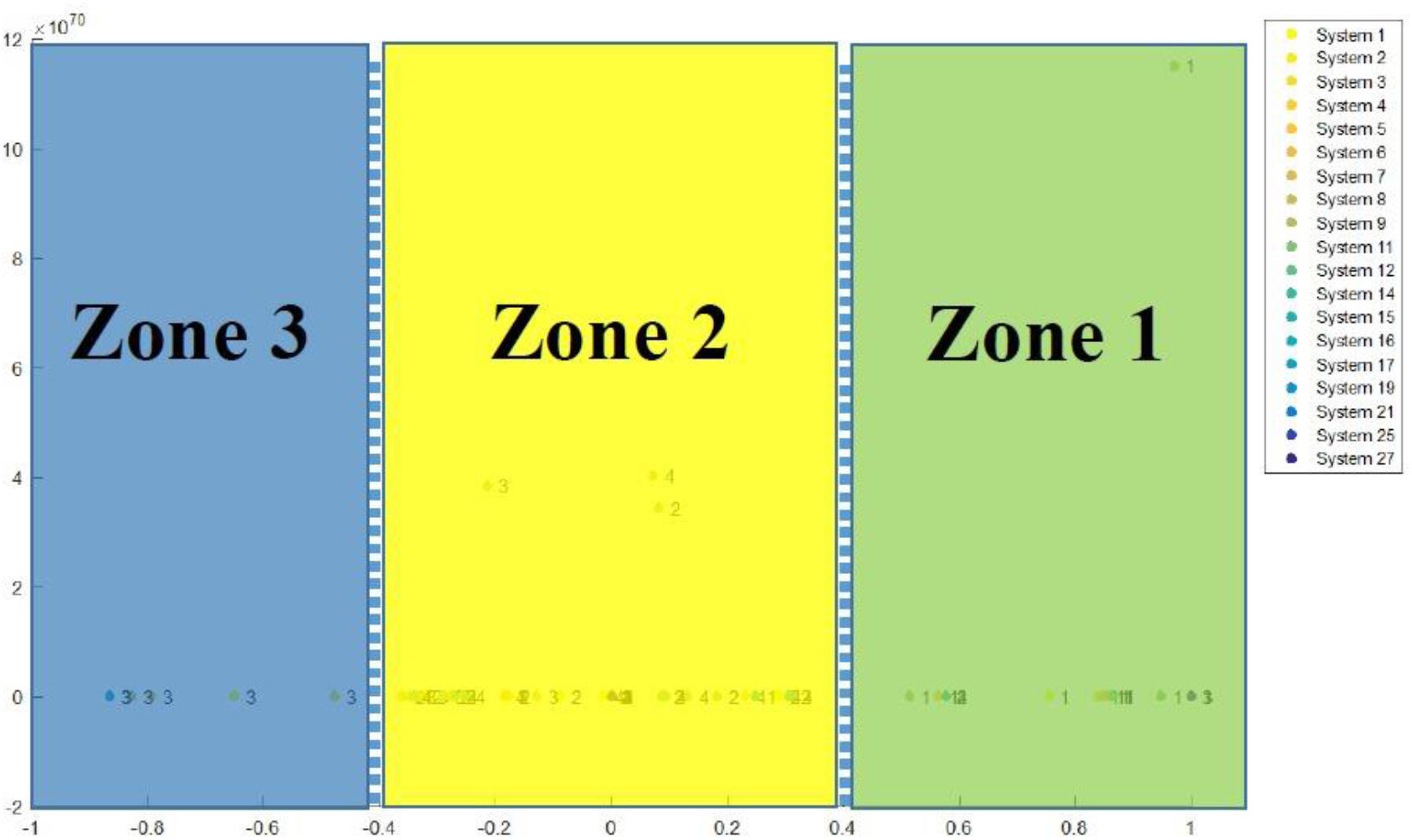
Zones 1, 2 and 3 for data 6th April 2020 to 2nd May 2020

## Analysis

One of the most significant results is the comparison with regard to the implementation of the FACS technique on the two sets of data is the alignment of zones. In Figure 0.8, these nodes occupied three quadrants of the plane that led to 3 identified zones with respect to data 28^th^ March 2020 – 5^th^ April 2020. On the other hand, these nodes occupied three sectors vertically for data 6^th^ April 2020 – 2^nd^ May 2020 (see Figure 0.12). The alignment of zones clearly indicates that the mitigation strategy taken by the Malaysian government has been fruitful and on the right track.

**Figure 0.1.**
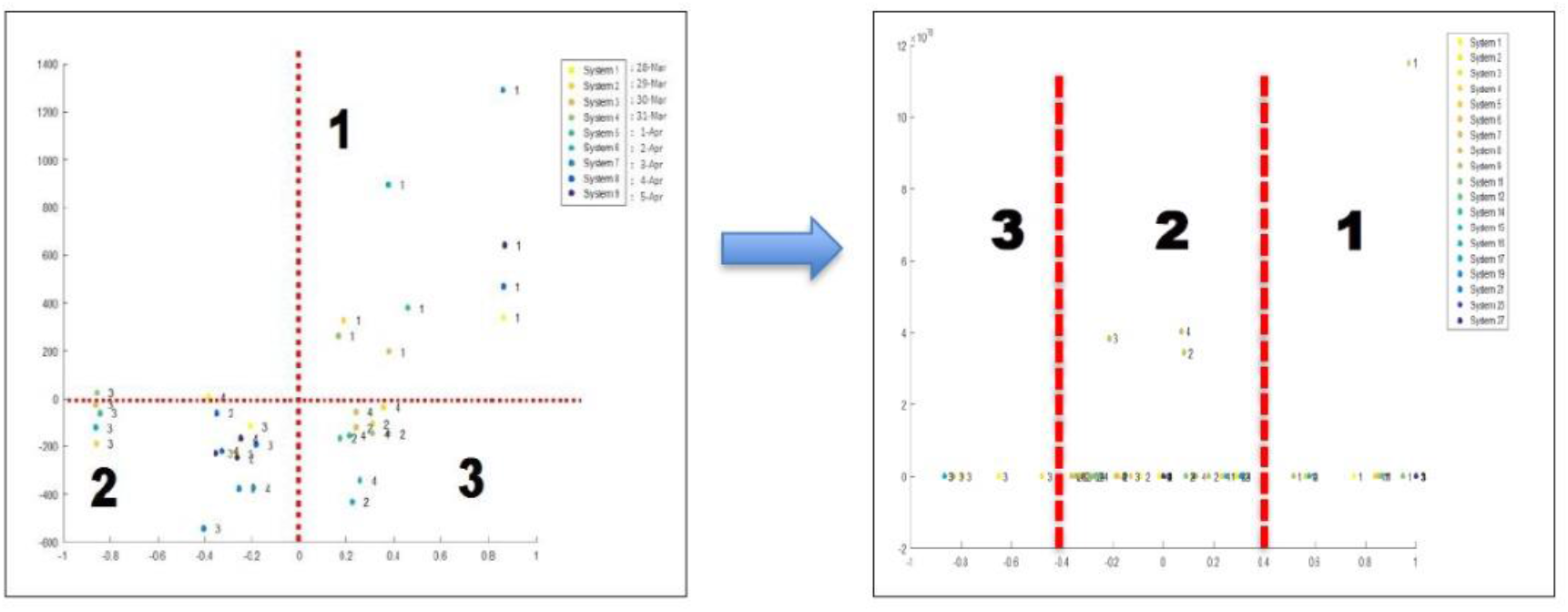
Alignment of zones.

## Conclusion

In this paper, we demonstrated a fuzzy autocatalytic analysis for the COVID-19 outbreak in Malaysia. The method is able to identify some significant features of the pandemic outbreak as well as some essential assessments on mitigation strategies. The method can be used to model any future pandemic.

### Data Availability

The data are obtained from the Ministry of Health (MOH) Malaysia and National Institutes of Health Malaysia (NIH) (publicly available).

### Conflicts of Interest

The authors declare that there is no conflict of interest regarding the publication of this paper.

## Acknowledgments

We gratefully acknowledge the Ministry of Health (MOH) and the National Institute of Health Malaysia for allowing us to use their published data (publicly available). We thank the Faculty of Science and Azman Hashim International Business School, Universiti Teknologi Malaysia, for their tremendous support for this work.

## Appendix

|  |  | Number of daily cases |  |  |  |  |  |  |  |  |  |  |  |  |  |  |  |  |  |  |  |  |  |  |  |  |  |  |
| --- | --- | --- | --- | --- | --- | --- | --- | --- | --- | --- | --- | --- | --- | --- | --- | --- | --- | --- | --- | --- | --- | --- | --- | --- | --- | --- | --- | --- |
| V | State | 6-Apr | 7-Apr | 8-Apr | 9-Apr | 10-Apr | 11-Apr | 12-Apr | 13-Apr | 14-Apr | 15-Apr | 16-Apr | 17-Apr | 18-Apr | 19-Apr | 20-Apr | 21-Apr | 22-Apr | 23-Apr | 24-Apr | 25-Apr | 26-Apr | 27-Apr | 28-Apr | 29-Apr | 30-Apr | 1-May | 2-May |
| 1 | Perlis | 0 | 4 | 0 | 0 | 0 | 0 | 0 | 0 | 1 | 0 | 0 | 0 | 0 | 0 | 0 | 0 | 0 | 0 | 0 | 0 | 0 | 0 | 0 | 0 | 0 | 0 | 0 |
| 2 | Kedah | 3 | 5 | 1 | 2 | 0 | 0 | 0 | 0 | 0 | 1 | 0 | 0 | 0 | 0 | 0 | 0 | 1 | 0 | 0 | 0 | 0 | 0 | 0 | 0 | 0 | 0 | 0 |
| 3 | Pulau Pinang | 0 | 1 | 6 | 0 | 1 | 5 | 2 | 0 | 3 | 0 | 0 | 0 | 0 | 0 | 0 | 0 | 0 | 0 | 0 | 0 | 2 | 0 | 0 | 0 | 0 | 0 | 0 |
| 4 | Perak | 6 | 14 | 4 | 4 | 1 | 1 | 1 | 3 | 3 | 0 | 1 | 1 | 0 | 0 | 0 | 0 | 0 | 0 | 0 | 0 | 0 | 1 | 0 | 0 | 0 | 0 | 0 |
| 5 | Selangor | 27 | 50 | 58 | 40 | 30 | 35 | 53 | 13 | 50 | 17 | 13 | 9 | 2 | 3 | 2 | 11 | 1 | 12 | 18 | 1 | 6 | 4 | 5 | 1 | 27 | 24 | 63 |
| 6 | Negeri Sembilan | 8 | 22 | 9 | 11 | 11 | 17 | 9 | 42 | 2 | 1 | 26 | 13 | 3 | 0 | 3 | 1 | 2 | 20 | 2 | 0 | 1 | 1 | 1 | 72 | 5 | 0 | 8 |
| 7 | Melaka | 10 | 4 | 13 | 3 | 14 | 8 | 3 | 0 | 3 | 0 | 0 | 1 | 0 | 37 | 2 | 1 | 1 | 0 | 3 | 0 | 1 | 0 | 0 | 0 | 5 | 1 | 2 |
| 8 | Johor | 30 | 10 | 18 | 7 | 20 | 21 | 14 | 21 | 8 | 14 | 13 | 9 | 16 | 3 | 2 | 1 | 3 | 3 | 4 | 0 | 4 | 0 | 1 | 0 | 3 | 3 | 1 |
| 9 | Pahang | 3 | 5 | 3 | 14 | 4 | 27 | 11 | 0 | 11 | 17 | 8 | 1 | 2 | 5 | 2 | 8 | 6 | 1 | 1 | 1 | 0 | 0 | 0 | 5 | 1 | 3 | 8 |
| 10 | Terengganu | 6 | 0 | 3 | 5 | 3 | 1 | 0 | 4 | 0 | 0 | 2 | 0 | 1 | 0 | 0 | 0 | 0 | 1 | 0 | 0 | 0 | 0 | 0 | 0 | 0 | 0 | 0 |
| 11 | Kelantan | 1 | 5 | 0 | 1 | 1 | 2 | 1 | 2 | 0 | 0 | 1 | 0 | 0 | 0 | 0 | 0 | 0 | 0 | 0 | 0 | 0 | 0 | 0 | 0 | 0 | 0 | 0 |
| 12 | Sabah | 2 | 1 | 7 | 0 | 2 | 10 | 6 | 14 | 5 | 0 | 3 | 5 | 10 | 3 | 2 | 0 | 0 | 1 | 2 | 0 | 0 | 1 | 1 | 2 | 0 | 0 | 0 |
| 13 | Sarawak | 12 | 15 | 18 | 6 | 15 | 7 | 8 | 4 | 15 | 4 | 16 | 10 | 6 | 5 | 6 | 22 | 19 | 4 | 9 | 10 | 7 | 3 | 8 | 6 | 5 | 2 | 9 |
| 14 | WP Kuala Lumpur | 18 | 31 | 14 | 14 | 14 | 46 | 44 | 27 | 69 | 27 | 26 | 19 | 14 | 19 | 4 | 13 | 16 | 29 | 49 | 39 | 16 | 30 | 14 | 8 | 10 | 32 | 14 |
| 15 | WP Labuan | 1 | 1 | 0 | 0 | 0 | 2 | 0 | 0 | 0 | 0 | 0 | 1 | 0 | 0 | 0 | 0 | 0 | 0 | 0 | 0 | 0 | 0 | 0 | 0 | 0 | 0 | 0 |
| 16 | WP Putrajaya | 4 | 2 | 2 | 2 | 2 | 2 | 1 | 2 | 0 | 0 | 1 | 0 | 0 | 9 | 13 | 0 | 1 | 0 | 0 | 0 | 1 | 0 | 1 | 0 | 1 | 4 | 0 |

## Notes

### Competing Interest Statement

The authors have declared no competing interest.

### Funding Statement

No external funding was recieved.

## References

[1] P. Zhou et al., “A pneumonia outbreak associated with a new coronavirus of probable bat origin,” Nature, vol. 579, no. 7798, pp. 270–273, 2020, doi: 10.1038/s41586-020-2012-7.

[2] K. Liang, “Mathematical model of infection kinetics and its analysis for COVID-19, SARS and MERS,” Infect. Genet. Evol., vol. 82, p. 104306, 2020, doi: https://doi.org/10.1016/j.meegid.2020.104306.

[3] Q. Lin et al., “A conceptual model for the coronavirus disease 2019 (COVID-19) outbreak in Wuhan, China with individual reaction and governmental action,” Int. J. Infect. Dis., vol. 93, pp. 211-216, Apr. 2020, doi: 10.1016/j.ijid.2020.02.058.

[4] S. G. Krantz and A. S. R. S. Rao, “Level of underreporting including underdiagnosis before the first peak of COVID-19 in various countries: Preliminary retrospective results based on wavelets and deterministic modeling,” Infect. Control Hosp. Epidemiol., pp. 1–3, 2020, doi: DOI: 10.1017/ice.2020.116.

[5] F. A. Binti Hamzah, C. Lau, H. Nazri, D. V Ligot, G. Lee, and C. L. Tan, “CoronaTracker: worldwide COVID-19 outbreak data analysis and prediction,” Bull World Heal. Organ. E-pub, vol. 19, 2020.

[6] W. G. Walter, “Slow potential waves in the human brain associated with expectancy, attention and decision,” Arch. Psychiatr. Nervenkr., vol. 206, no. 3, pp. 309–322, 1964, doi: 10.1007/BF00341700.

[7] J. Jia, J. Ding, S. Liu, G. Liao, and J. Li, “Modeling the control of COVID-19: impact of policy interventions and meteorological factors,” Electron. J. Differ. Equations, vol. 2020, pp. 1–24, 2020.

[8] P. Forster, L. Forster, C. Renfrew, and M. Forster, “Phylogenetic network analysis of SARS-CoV-2 genomes,” Proc. Natl. Acad. Sci., vol. 117, no. 17, pp. 9241 LP—9243, Apr. 2020, doi: 10.1073/pnas.2004999117.

[9] S. S. Mamat, S. R. Awang, and T. Ahmad, “Preference Graph of Potential Method as a Fuzzy Graph,” Adv. Fuzzy Syst., vol. 2020, p. 8697890, 2020, doi: 10.1155/2020/8697890.

[10] A. Rosenfeld, “Fuzzy graphs, Fuzzy Sets and their Applications.” Academic Press, New York, 1975.

[11] S. A. Kauffman, “Cellular Homeostasis, Epigenesis and Replication in Randomly Aggregated Macromolecular Systems,” J. Cybern., vol. 1, no. 1, pp. 71-96, Jan. 1971, doi: 10.1080/01969727108545830.283.

[12] O. E. Rössler, “Ein systemtheoretisches Modell zur Biogenese / A System Theoretic Model of 284 Biogenesis,” Zeitschrift für Naturforsch. B, vol. 26, no. 8, pp. 741–746, 1971, doi: https://doi.org/10.1515/znb-1971-0801.

[13] S. Jain and S. Krishna, “Autocatalytic sets and the growth of complexity in an evolutionary model,” 287 Phys. Rev. Lett., vol. 81, no. 25, p. 5684, 1998.

[14] T. Ahmad, S. Baharun, and K. A. Arshad, “Modeling a clinical incineration process using fuzzy 289 autocatalytic set,” J. Math. Chem., vol. 47, no. 4, pp. 1263–1273, 2010, doi: 10.1007/s10910-009-9650-1.

[15] A. Ashaari, T. Ahmad, S. Zenian, and N. A. Shukor, “Selection probe of EEG using dynamic graph of autocatalytic set (ACS),” in Communications in Computer and Information Science, 2016, vol. 652, pp. 293 25–36, doi: 10.1007/978-981-10-2777-2_3.

[16] N. Hassan, T. Ahmad, and N. Mohd Zain, “A novel chemometrics fuzzy autocatalytic set (FACS) with fourier transform infrared (FTIR) spectroscopy for halal authentication of gelatins,” Sains Malaysiana 296 (In Press., 2020.

[17] T. Ahmad, S. A. Bakar, S. Baharun, and F. A. M. Binjadhnan, “Coordinated transformation for fuzzy autocatalytic set of fuzzy graph type-3,” J. Math. Stat., vol. 11, no. 4, pp. 119–127, 2016, doi: 10.3844/jmssp.2015.119.127.

